# Diagnostic evaluation of the presence of residues of glyphosate-AMPA and 2.4D pesticides in urine samples from people living in a rural Brazilian community

**DOI:** 10.1101/2021.08.16.21259798

**Authors:** Luciano Z. P. Candiotto, Mariane Okamoto Ferreira, Isadora Nunes Ferreira, Géssica Tuani Teixeira, Janaína Carla da Silva, Emanueli Hammes Tedesco, Shaiane Carla Gaboardi, Carolina Panis

## Abstract

Pesticide contamination in rural communities is a known problem worldwide. In this study, we investigated the levels of pesticide residues in urine samples from residents of a rural community located in the municipality of Francisco Beltrão-Paraná, Brazil. According to the residents’ report obtained through a structured interview, the main form of exposure has been due to the drift of pesticides resulting from the spraying carried out on crops neighboring the community, with one crop existing on the left edge and another on the right edge of the site. The investigation was based on a demand from residents concerned about the frequent occurrence of symptoms after spraying on neighboring crops, such as nausea, headaches, and dizziness. Thus, samples were collected immediately after the occurrence of spraying in the crop surrounding the site. In the collected samples (n=35, 1 sample per family) laboratory analyzes were performed to identify possible pesticide residues. To identify possible contaminating pesticides in the samples, multi-residue analysis was performed by gas chromatography coupled with mass spectrometry (GC-MS/MS). To identify 2.4D residues, an active widely used in the region and not detectable by the technique used for other pesticides, the analyzes were performed using the enzyme immunoassay technique. The results indicate that all urine samples collected showed the presence of 2.4D residues and that 90% of them presented the presence of glyphosate-AMPA residues, without the detection of the other investigated residues. The analysis of data obtained from interviews with residents shows an association between living in the place for longer and having cases of abortion (p<0.05, R=0.43) or feeling unwell after the application of the poison by the neighboring crop (p<0.05, R=0.47). In addition, residents who reported being poisoned with poison also reported having cancer (p<0.05, R=0.42). Assuming that it is unacceptable that urine samples have pesticide residues, it is concluded that the residents of this community are widely contaminated by pesticides potentially coming from the spraying of the neighboring crop, especially by pesticides that have the active ingredients in their formulas. 2,4D and glyphosate-AMPA and that may be impacting the health of these people in the long term.

## Introduction

The current scenario of agricultural production indicates an increasing use of chemical compounds, called pesticides, under the argument of maximizing productivity rates and, consequently, profit from agricultural activities. In Brazil, in the last decade, the consumption of legally commercialized pesticides rose from 305 thousand tons in 2009 to about 540 thousand tons in 2017 (Gaboardi et al., 2019).

This expansion in the marketing and use of pesticides, in addition to economic “benefits”, has been responsible for several environmental (and social) impacts, mainly linked to the contamination of water, food, soil, animals and human beings, as pointed out by Dossier of the Brazilian Association of Public Health (ABRASCO) (Carneiro et al., 2015), and by other studies and publications (Gaboardi, 2021; Pignati, 2007; Porto, 2007 and 2018; Thomaz Júnior, 2017; Bombardi, 2007, 2011 and 2013).

The negative consequences of the use of pesticides have been the subject of debates in the scientific and political sphere, so that the topic involves controversies and questions that require objective and precise answers. On the one hand, sectors linked to agribusiness defend the use of these products, claiming that it is not possible to feed the world population without them. On the other hand, institutions linked to collective health and environmental protection have questioned the need and effectiveness of pesticides, given their negative effects on ecosystems and humanity.

Concern about the consumption of pesticides and their consequences has grown in Brazil, considering that the country currently figures on the international scene, along with China and the United States, as one of the leaders in the sale of pesticides in the world (FAOSTAT, 2021). Furthermore, the massive use of active ingredients occurs, mainly, in states specialized in the production of agricultural commodities, such as Paraná, which is the second largest grain producer in Brazil, especially soy, corn, wheat and beans.

This specialized production, dependent on the use of pesticides, makes Paraná the fourth largest marketer of active ingredients in Brazil, and this reality is similar in all mesoregions where agribusiness is present. In the Southwest of Paraná, approximately seven thousand and five hundred tons of pesticides have been sold per year, which are mainly destined for soybean and corn monocultures.

In the municipality of Francisco Beltrão, one of the municipalities that most commercialize pesticides in the Southwestern Paraná mesoregion, between 2011 and 2019, an annual average of 11 kilos of pesticides per hectare of cultivated area was used, according to data reported in the Trade and Monitoring System. Use of Pesticides in Paraná (SIAGRO), linked to the Agricultural Defense Agency of Paraná (ADAPAR), and disclosed by the Paraná Institute for Economic and Social Development (IPARDES). This annual average is higher than that of the state of Paraná and Brazil, which are, respectively, 5.4 kg/ha and 6.7 kg/ha (IBGE, 2014).

Despite the widespread use of these products in Brazil, scientific evidence is needed to qualify the debate on the environmental and social consequences of this use (Gaboardi et al., 2019). Therefore, the objective of this project was to investigate the presence of pesticide residues in urine samples from residents of a community located in the rural region of the municipality of Francisco Beltrão-PR after an extensive pesticide pulverization in a large neighbor crop.

## METHODS

### Ethical aspects

The study is approved by the Institutional Ethics Committee from the State University of West Paraná and is available for public consultation under number 810.501 and CAAE 35524814.4.0000.0107.

### Study location

The study area comprises a rural community located in the municipality of Francisco Beltrão, Paraná, Brazil (Fig. 1), composed of about 50 families (140 people), living in plots of about 5,000 m2. This population lives adjacent to agricultural areas where pesticide spraying in large quantities by third parties is frequent (crops bordering the community), as shown in Fig. 2. Crops with more intense pesticide spraying are located to the west and east of the community. To the north, there is a plantation of eucalyptus (Eucalyptus) and to the south, another crop.

**Figure 1:**
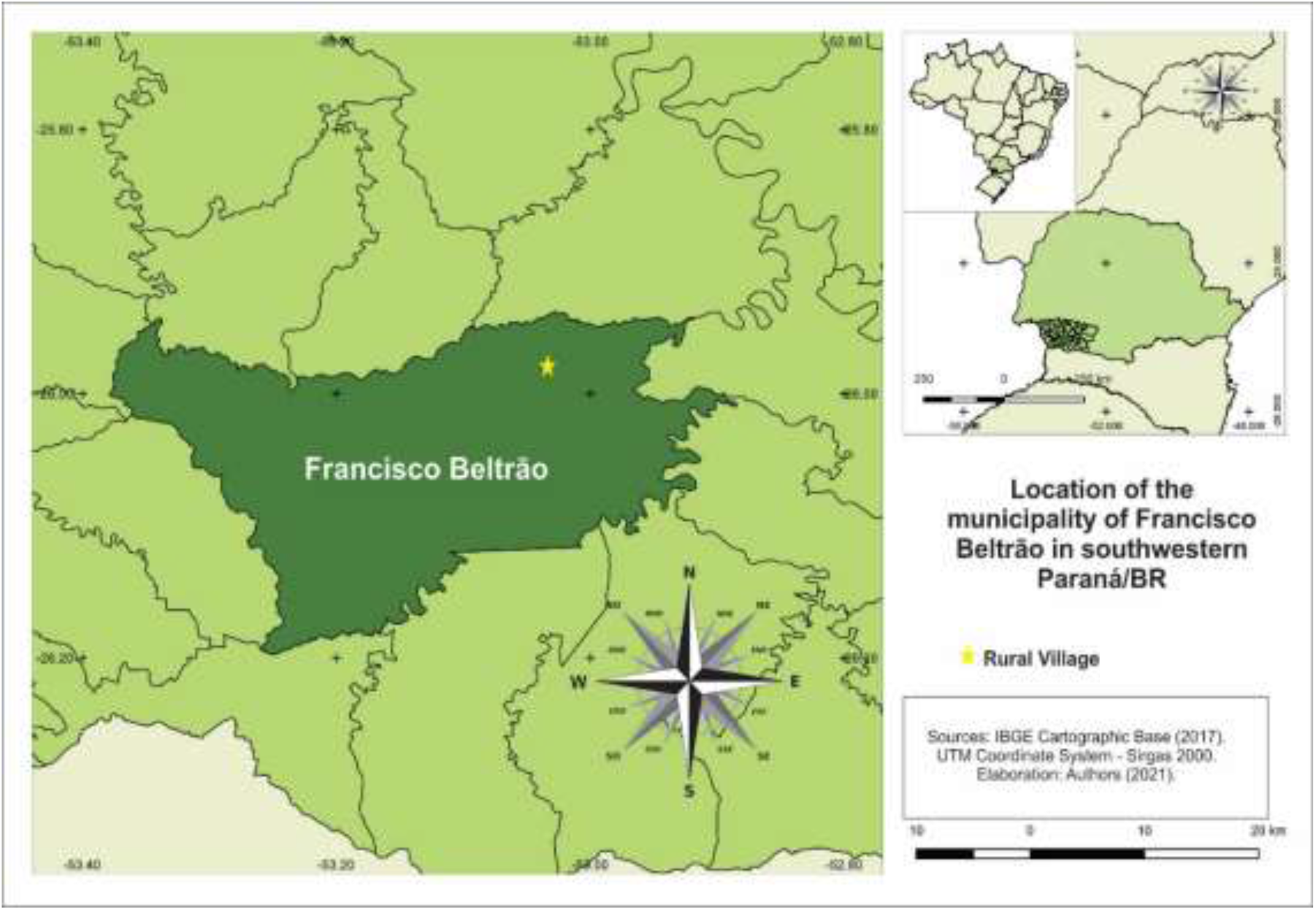
Location map of the study community, in Francisco Beltrão, Paraná, Brazil

**Figure 2.**
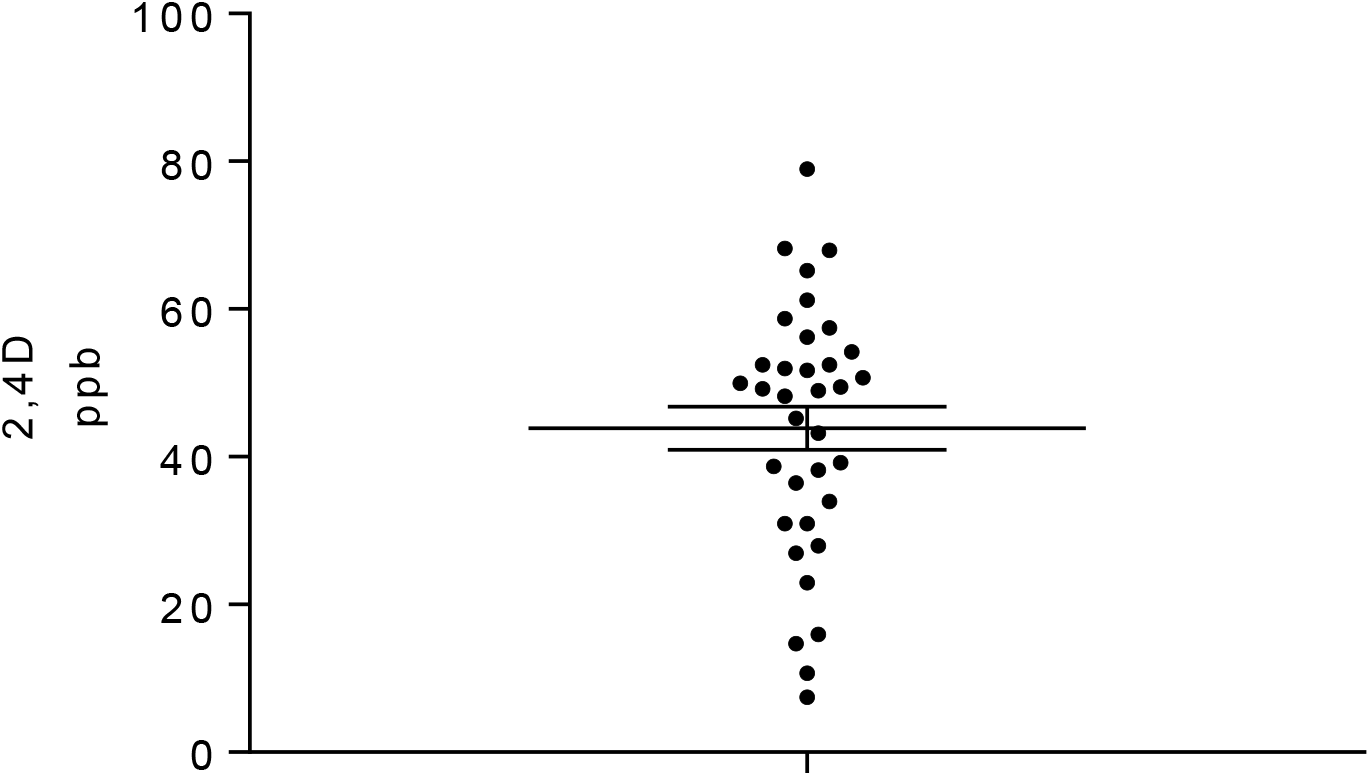
2.4D levels detected in the urine samples of the studied population.

The crops bordering the community (west, east and south) have intercropped soybean, corn, wheat and beans. In the year 2021, the summer crop of the western crop was occupied by soybeans, followed by corn. The last planting was edible beans, harvested in June 2021. The crop to the east, on the other hand, had the 2021 summer crop occupied by soybeans, followed by the planting of wheat, which currently occupies the agricultural area. These two crops are managed by tenants, that is, by people who rent the land to plant. Both are owned by the same family. In turn, tenants usually hire third parties to prepare the soil, plant, manage (spray during plant development) and harvest agricultural products.

The crop in the south of the community is managed by the owner, who has been planting corn and soybeans. However, as it has a smaller boundary area with the studied community than the west and east crops, the impact of spraying on this crop is smaller.

### Sample collection and analysis of pesticide levels

After signing the free and informed consent form, urine samples were collected from 35 families, and the resident who stayed longer in the household during spraying was selected as the sample donor for analysis. This collection was carried out at an interval within a maximum of 6 hours after spraying pesticides on the neighboring field, in order to preserve the detection of pesticides with a reduced half-life.

To determine the levels of pesticides, the collected samples were analyzed by gas chromatography coupled with mass spectrometry using the multi-residue pesticide analysis technique (GC-MS/MS), with extraction based on a protocol previously published by Maffei, Nogueira and Brondi (10). The following running conditions were used: column (ChemElut S-Agilent techologies), preactivated with acetonitrile, pressure 2 to 3 psi, analyzed by gas chromatography coupled to a mass spectrometry detector, according to the EPA 8270 method. The analysis allows the detection of 55 different pesticides, listed in Supplementary File 2. As the GC-MS/MS did not allow the detection of 2.4D, one of the main actives sprayed on the region’s crops, a commercial enzyme immunoassay kit (Abraxis LLC) was used ^™^, Warminster, Pennsylvania, USA) for this analysis. The test’s limit of quantification is 1.67 ppb.

### Epidemiological data

A data collection instrument (Supplementary File 1) was applied to identify the profile of exposure to pesticides based on three main elements: 1) exposure to pesticides in the present; 2) exposure to pesticides in the past and; 3) general aspects of health and intoxication. The questions of the data collection instrument allowed the identification of information regarding the municipality of residence, time the individual lived or lives in the rural area, identification of agricultural production with the use of pesticides, as well as whether the individual has applied or applied any type of pesticide throughout life, and for how long. In addition, it was possible to identify whether the individual had contact with pesticides by washing clothes and decontaminating Personal Protective Equipment (PPE), as well as whether PPE was used during pesticide spraying. It was also possible to identify by the instrument where the water that residents consume comes from, as well as the distance of this water from the farming areas. Regarding health aspects, respondents were asked about the occurrence of acute poisoning by pesticides, and about the presence of the following conditions (in him and in his family): diagnosis of cancer, cases of malformation, abortion, difficulty in getting pregnant, as well as of exposure to pesticides before the children’s conception.

### Statistical analysis

Questionnaire data and pesticide analysis results were tabulated in Excel and correlations analyzed using SPSS 24.0 (IBM, USA) and GraphPad Prism 7.0 software.

## RESULTS

### Questionnaire analysis

The questionnaires applied to the study population aimed to provide an overview of the history of rural life and the direct and indirect exposure of residents to pesticides in their lives; the eventual use and current contact with pesticides by them (use or drift); events related to poisoning and other health problems that have been associated with contact with pesticides (cancer, abortion, malformation), among other factors.

Questionnaires were applied to 33 residents who were present and agreed to answer them, 2 were not at the time of collections. Considering the central question of the study (about the possible association between exposure to applied pesticides and health problems for residents), the study of data collected from the residents of Vila Rural indicates the existence of the following correlations:

1. Those who have lived in the place longer have a history of miscarriage (Q 51 × Q 31), R=0.43;
2. Residents who report feeling sick after their neighbors apply poison also report that they have already been poisoned by poison (Q 22 × Q 27), R=0.47;
3. Residents who report having already been intoxicated with poison also report having cancer (Q 26 × Q 28), R=0.42;
4. Specifically, residents who reside on the margins of neighboring crops, in direct contact with the crop, reported that:

a. Does not use poison today (Q 34 × Q 3), R=-0.44;
b. Does not apply any type of poison, neither in crops nor at home (Q 34 × Q 5), R=-0.58;
c. Believes that the poison applied to the neighboring crop is bad for your health (Q 34 × Q 22), R=0.66.

### Pesticide levels

It was not possible to know exactly which pesticides and active ingredients were used in the crops neighboring the study community. However, it was possible to verify that in the pulverized property there is the current use of glyphosate, 2.4D and atrazine, according to information provided by the Agricultural Defense Agency of Paraná (ADAPAR), unit of Francisco Beltrão. Considering the widespread use of glyphosate and 2.4 D in the state of Paraná (35% of total pesticides sold in 2019), as well as the predominance of soybean cultivation in the state (55.5% of the harvested area), it is possible estimate that these two active ingredients have possibly been used in crops bordering the community studied at the time of collection of the analyzed samples.

In this context, the results of the analysis of urine indicate that the studied population was exposed and severely contaminated by 2,4D and glyphosate-AMPA residues. The results of these analyzes are shown in Figures 2 and 3. Although multiresidue analyzes were performed, which allow the identification of the presence of several active ingredients, the only active ingredient found in the samples was glyphosate and its main metabolite, AMPA. The presence of 2,4D was also detected in all samples tested using the enzyme immunoassay kit.

**Figure 3.**
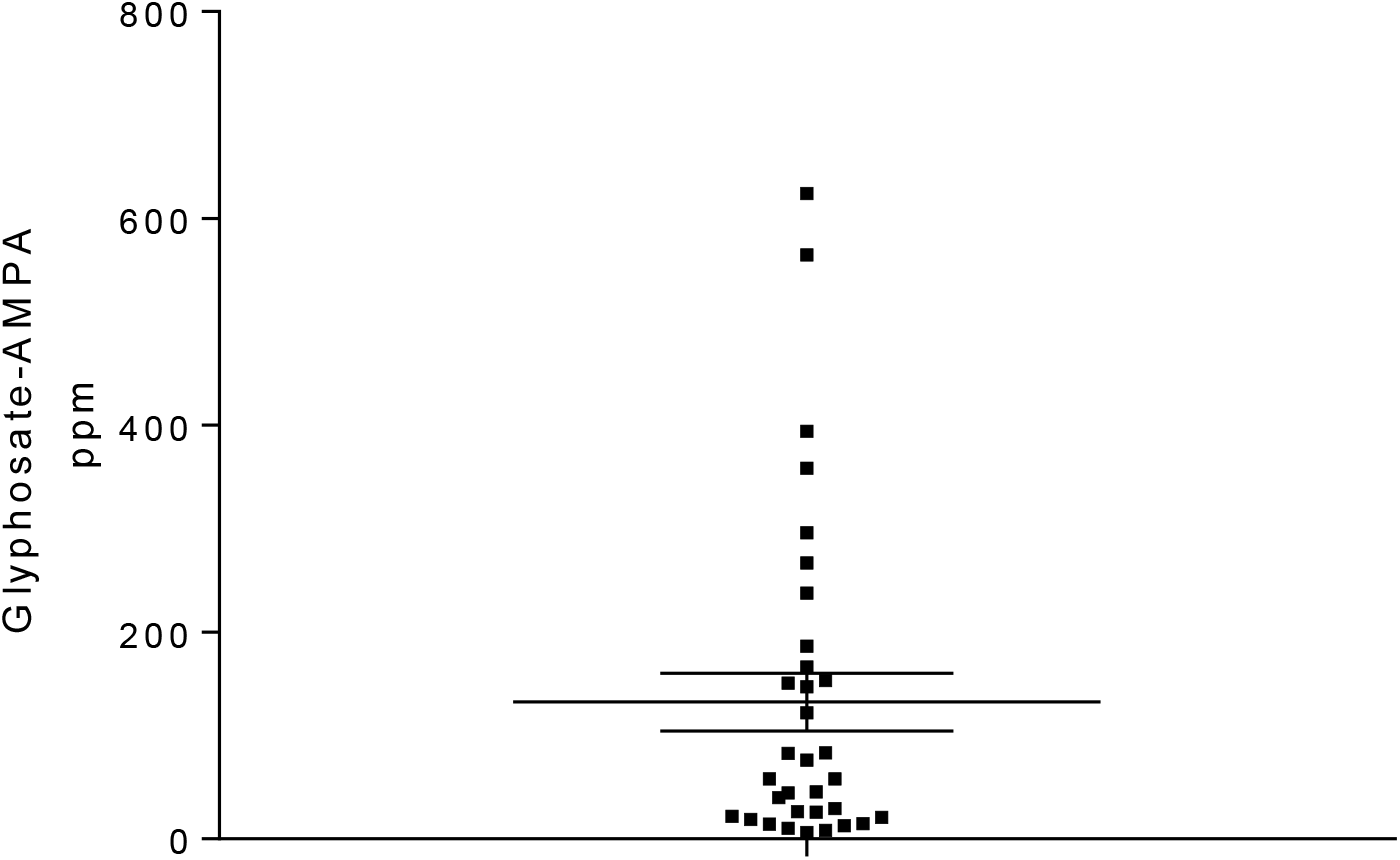
Glyphosate-AMPA levels detected in the urine samples of the studied population.

Of a total of 35 urine samples collected and analyzed, 24 were from women and 11 from men. Regarding urine samples, it is assumed that no pesticide residues should be found in any of the samples. However, all 35 samples collected had 2.4 D residues, ranging from 7.4 to 78.9 ppb. When separating the results into classes, it can be seen that: 1) 4 samples (11.4%) presented values from 0 to 20 ppb; 2) 10 samples (28.6%) had values from 21 to 40 ppb; 3) 16 samples (45.7%) presented values from 41 to 60 ppb; and 4) 5 samples (14.3%) presented values from 61 to 80 ppb.

The results for glyphosate-AMPA showed more expressive data when compared to the results for 2.4 D. Of the 35 urine samples, only two were not identified with glyphosate residues, however, the maximum value identified was 624,327 ppb (women). When separating the results into classes, it can be seen that: 1) 20 samples (57.1%) presented values from 6,000 to 100,000 ppb; 2) 6 samples (17.1%) presented values from 101,000 to 200,000 ppb; 3) 3 samples (8.6%) had values from 201,000 to 300,000 ppb; and 4) 4 samples (11.4%) presented values above 300,000 ppb.

## DISCUSSION

This study records the occurrence of extensive human contamination by spraying pesticides, which resulted in the identification of elevated levels of 2,4D and glyphosate-AMPA in urine samples from exposed individuals. Considering the short half-life of glyphosate (approximately 6 hours) and the stability of 2.4D for longer periods, the levels of pesticides found suggest that glyphosate contamination probably occurred very close to the date of collection of urine samples, while 2.4D levels suggest contamination that may be current or past (weeks). There is no knowledge of another source capable of resulting in as high concentrations of glyphosate in urine as detected.

The two active ingredients found in the urine samples of the studied population are herbicides widely used in Brazil, mainly in soybean and corn crops. Glyphosate (N-(phosphonomethyl)glycine) is a broad-spectrum, systemic herbicide that suppresses the growth of plants such as grasses, perennials, shrubs and trees. Currently, it is the most popular and handled pesticide in the world, registered in approximately 130 countries (Meftaul et al., 2020). It can be used for agricultural and non-agricultural purposes, such as in gardening, on roadsides and in industrial yards. 2,4-D, (2,4-dichlorophenoxy) acetic acid, is also a systemic action herbicide, used to control weeds in soybean, corn, wheat, rice, etc. crops. It is among the most widely distributed pollutants in the environment (Islam et al., 2018). This active ingredient is present in low concentration in superficial areas of regions where its use is high and the highest concentrations are detected in the soil, air and in surface waters surrounded by cultivated fields.

Glyphosate and 2,4-D are the two most commercialized active ingredients in Brazil. In 2019, 217,592.24 tons of glyphosate and 52,426.92 tons of 2,4-D were sold, which represented 43.5% of the total sales volume of pesticides in the country (IBAMA, 2019). In Paraná, in 2019, Glyphosate was also the most commercialized active ingredient, followed by Paraquate and 2,4-D. Thus, 27,622.38 tons of glyphosate and 6,074.33 tons of 2,4-D and its salts were sold, which represents 35.3% of the total volume of pesticides sold in the state that year (ADAPAR, 2019; Gaboardi, 2021).

The wide use of these active ingredients has raised many questions and concerns about their possible effects on human health. This is because Glyphosate and 2,4-D have been classified by the IARC of the World Health Organization as products likely and possibly carcinogenic to humans (Group 2A and Group 2B, respectively) (IARC, 2015; IARC, 2018). However, both continue to be widely marketed in Brazil and commonly used in soybean, corn and wheat crops in Paraná and in the southwestern region of the state, where the municipality of Francisco Beltrão and the community studied are located.

Studies show that several correlations have been found between the use of glyphosate and the occurrence of diseases, including various forms of cancer (Fortes et al., 2016), kidney damage (Jayasumana et al., 2014) and neurological problems such as autism (Fluegge and Fluegge, 2016), Alzheimer’s and Parkinson’s disease (Swanson et al., 2014). Miscarriages and dermatological and respiratory diseases were also related to exposure to glyphosate (Camacho and Mejía, 2017). Likewise, 2,4-D has been associated with several health problems, such as childhood cancer (Flower et al., 2003), prostate cancer (Koutros et al., 2012) and melanoma (Dennis et al., 2010). Furthermore, there is evidence that 2,4-D is related to oxidative stress (Lerro et al., 2017) and immunosuppression (IARC, 2018). A study published by Tan et. al. (2016) suggested that exposure to 2,4-D and its accumulation in seminal plasma and follicular fluid may increase the risk of male infertility. of the residents of Vila Rural Água Viva, municipality of Francisco Beltrão/PR, is worrisome, given the health risks possibly associated with these pesticides. The active ingredients glyphosate and 2,4-D were detected in 94.2% and 100% of the samples (n=35), respectively.

A survey conducted in ten municipalities in the Southwest of Paraná (Gaboardi et al., 2021) found contamination in urine samples by glyphosate in 58.3% of the cases evaluated, with a mean value of 1.14 ppb. 2,4-D was detected in 34.7% of the samples analyzed, with an average value of 58.11 ppb. In another study, glyphosate was detected in the breast milk of 100% of samples collected in the municipality of Francisco Beltrão, at the peak of spraying of corn and soybeans in 2018, recording average values of 1.45 ppb. The results suggest the possibility of contamination by Glyphosate in the lactating population studied, indirectly, through water, contaminated food and air, considering that the agricultural production process adopted in the municipality of Francisco Beltrão/PR includes the intensive use of pesticides in crops (Camiccia, 2019).

Other researches have reported the presence of glyphosate in biological samples in Brazil. In a study carried out in the state of Mato Grosso, 79 residents of rural and urban areas in the municipality of Lucas do Rio Verde were selected and 88% of positive samples were observed. Among the samples from the rural population (n=42), 83% were positive for glyphosate with values detected between 0.38 ppb and 5.05 ppb, based on the methodology of colorimetric kits with ELISA reading (Belo et al., 2012). This same analysis technique was applied in another research conducted in Santarém, in the western region of Pará, where there is an expansion of the national agricultural frontier towards the Amazon. Twenty-seven urine samples were collected from rural communities in the metropolitan region of Santarém, and residual concentrations of glyphosate ranging from 0.31 to 4 ppb were detected (Schwamborn, 2019).

Another study carried out in the state of Mato Grosso, in the Nova Mutum region, analyzed 90 urine samples from farmers between 2017 and 2018 and 12% of these samples had glyphosate levels. The methodology used in the analyzes was performed by HPLC-FL and the detected values ranged from 0.34 and 1.15 ppb (Melo et al., 2020). Glyphosate has been detected in urine samples from people exposed to this active ingredient in countries other than Brazil (Cannolly et al., 2020). However, the values found seem to be much lower than the results identified in the urine analysis of the studied community residents. These results indicate that the contamination found in our study is quite high. These differences can be explained by the immediate collection after spraying, minimizing the degradation of the studied assets, and by the high detection capacity of the GC-MS/MS technique compared to other methodologies.

The glyphosate levels found in our study could potentially cause some type of intoxication, even subclinical, with chronic consequences to the health in the future. Zouaoui et al. (2013) reported very high values of glyphosate in the urine in patients with acute intoxication and with mild to moderate, severe and fatal symptoms. The researchers detected by chromatography maximum values of 3×10^6^ ppb in mild cases, 21.1×10^6^ ppb in cases of intoxication with severe symptoms and 22.3×10^6^ ppb in fatal cases. Another recent study carried out in China (Zhang et al., 2020) also detected a result very close to our findings in workers from glyphosate manufacturing industries, with a maximum detected value of 17,000 ppb.

Occupational exposure studies indicate lower glyphosate values. In France, Mesnage et al., 2012 found a maximum concentration value of 9.5 ppb in the urine of a pesticide applicator 7 hours after the start of handling glyphosate. In Ireland, Connolly et al., 2017 analyzed urine samples from 40 horticulturists before and after work activities and found that urine concentrations increased significantly post-work and had a geometric mean of 0.66 ppb glyphosate concentration, with a maximum value of 10 ppb. In Mexico, Osten et al., 2017, also detected glyphosate in the urine of a group of 81 subsistence farmers in the state of Campeche, at an average concentration of 0.47 ppb. In the United States, Perry et al., 2019 tested 18 cryopreserved samples from farmers who reported application of glyphosate in the 8 hours prior to sample collection and 39% showed detectable levels of glyphosate with an average concentration of 4.04 ppb, with the level maximum reached 12.0 ppb.

Regarding 2.4D, the Agricultural Health Study (AHS) reported its detection in urine samples in a group of 69 pesticide applicators from North Carolina and Iowa, with mean values of 7.8 and 25 ppb pre and post application, respectively, and the analyzes were performed by gas chromatography/mass spectrometry (GC / MS) (Thomas et al., 2010). In Ohio and North Carolina, another study, using the same analysis technique, investigated the exposure to 2,4-D of 135 children and their adult caregivers, and the active ingredient was detected in more than 85% of urine samples of children and adults in both states (Morgan et. al., 2008). Another study carried out in Poland conducted by Jurewicz et al. (2012), indicated that farmers’ wives can be exposed to 2,4-D, even if they do not participate in the spraying process. That’s because analyzes of urine samples from a group of 24 women were tested and showed residues of this pesticide at average concentrations of 3 ppb in the morning before spraying to 7.9 ppb the morning after spraying 2,4-D by their spouses.

Based on the results of the literature, it is observed that the results of the analyzes carried out on the samples of this study show a process of environmental injustice (Acselrad, 2010; Souza, 2019; Candiotto, 2021; Gaboardi, 2021), resulting from the widespread use of pesticides. In Brazil, especially, there is a process of human and environmental contamination throughout the country. Despite the difficulties in conducting research on this contamination, the increasing release of new pesticides in Brazil (Fig. 4) in recent years indicates that there is a trend of increasing contamination by pesticides in the country.

**Figure 4.**
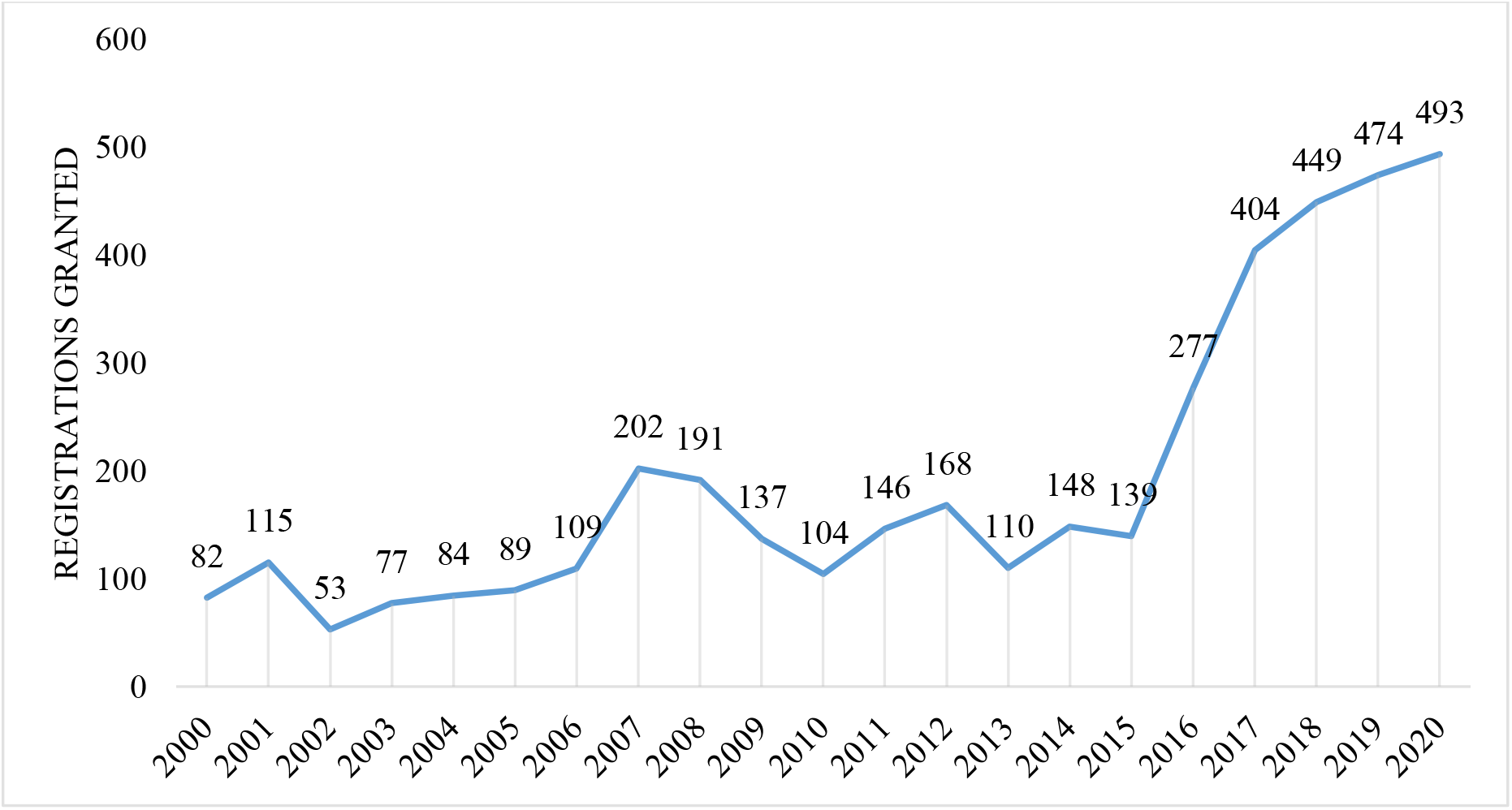
Total of pesticides registered per year in Brazil from 2000 to 2020.

Thus, it is also possible to affirm that the Brazilian State, with emphasis on the current federal government, but also including state and municipal governments, has promoted and/or colluded with a necropolitics (Mbembe, 2011 and 2018; Mondardo, 2019).

## CONCLUSIONS

The results obtained demonstrate extensive contamination of the population studied by pesticides 2.4D and glyphosate-AMPA, possibly by the drift of pesticides applied in crops bordering the community. Considering that almost all analyzed urine samples showed residues of glyphosate and 2.4 D, which are used in corn, soybean and wheat crops in the neighboring Vila Rural, it is suggested that part of this exposure is possibly linked to the drift of spraying carried out on neighboring crops. To minimize the problem, actions are needed to reduce pesticide drift from these crops, such as planting vegetable barriers around the community, and warning residents about when spraying will occur, among other legal and ethical measures to protect these people.

## Data Availability

All data are shown in the manuscript.

