## Supplementary file 1 for "Diagnostic evaluation of the presence of residues of glyphosate-AMPA and 2.4D pesticides in urine samples from people living in a rural Brazilian community"

**PATIENT IDENTIFICATION**

| NOME | Date of birth __/__/___ | Gender: ( ) F ( )M |
| --- | --- | --- |
| ADDRESS | | |
| MUNICIPALITY | PHONE number: | |

**FAMILY IDENTIFICATION**

| PATIENT name: | AGE | JOB |
| --- | --- | --- |
|  | 1 \| \| 0-10 \| 7 \| \| 61-70; 2 \| \| 11-20 \| 8 \| \| + 70; 3 \| \| 21-30 \| \| \|; 4 \| \| 31-40 \| \| \|; 5 \| \| 41-50 \| \| \|; 6 \| \| 51-60 \| \| \| | 1 \| \| Rural Worker; 2 \| \| Urban Worker; 3 \| \| Student; 4 \| \| Housewife; 5 \| \| Retired rural worker; 6 \| \| Retired urban worker |
| RELATIVE 1 (Name and relative type) | AGE | JOB |
|  | 1 \| \| 0-10 \| 7 \| \| 61-70; 2 \| \| 11-20 \| 8 \| \| + 70; 3 \| \| 21-30 \| \| \|; 4 \| \| 31-40 \| \| \|; 5 \| \| 41-50 \| \| \|; 6 \| \| 51-60 \| \| \| | 1 \| \| Rural Worker; 2 \| \| Urban Worker; 3 \| \| Student; 4 \| \| Housewife; 5 \| \| Retired rural worker; 6 \| \| Retired urban worker |
| RELATIVE 2 (Name and relative type) | AGE | JOB |
|  | 1 \| \| 0-10 \| 7 \| \| 61-70; 2 \| \| 11-20 \| 8 \| \| + 70; 3 \| \| 21-30 \| \| \|; 4 \| \| 31-40 \| \| \|; 5 \| \| 41-50 \| \| \|; 6 \| \| 51-60 \| \| \| | 1 \| \| Rural Worker; 2 \| \| Urban Worker; 3 \| \| Student; 4 \| \| Housewife; 5 \| \| Retired rural worker; 6 \| \| Retired urban worker |
| RELATIVE 3 (Name and relative type) | AGE | JOB |
|  | 1 \| \| 0-10 \| 7 \| \| 61-70; 2 \| \| 11-20 \| 8 \| \| + 70; 3 \| \| 21-30 \| \| \|; 4 \| \| 31-40 \| \| \|; 5 \| \| 41-50 \| \| \|; 6 \| \| 51-60 \| \| \| | 1 \| \| Rural Worker; 2 \| \| Urban Worker; 3 \| \| Student; 4 \| \| Housewife; 5 \| \| Retired rural worker; 6 \| \| Retired urban worker |
| RELATIVE 4 (Name and relative type) | AGE | JOB |
|  | 1 \| \| 0-10 \| 7 \| \| 61-70; 2 \| \| 11-20 \| 8 \| \| + 70; 3 \| \| 21-30 \| \| \|; 4 \| \| 31-40 \| \| \|; 5 \| \| 41-50 \| \| \|; 6 \| \| 51-60 \| \| \| | 1 \| \| Rural Worker; 2 \| \| Urban Worker; 3 \| \| Student; 4 \| \| Housewife; 5 \| \| Retired rural worker; 6 \| \| Retired urban worker |
| RELATIVE 5 (Name and relative type) | AGE | JOB |
|  | 1 \| \| 0-10 \| 7 \| \| 61-70; 2 \| \| 11-20 \| 8 \| \| + 70; 3 \| \| 21-30 \| \| \|; 4 \| \| 31-40 \| \| \|; 5 \| \| 41-50 \| \| \|; 6 \| \| 51-60 \| \| \| | 1 \| \| Rural Worker; 2 \| \| Urban Worker; 3 \| \| Student; 4 \| \| Housewife; 5 \| \| Retired rural worker; 6 \| \| Retired urban worker |

1. **Which municipality do you live?**

| 1 |  | Ampére | 12 |  | Manfrinópolis | 23 |  | Salto do Lontra |
| --- | --- | --- | --- | --- | --- | --- | --- | --- |
| 2 |  | Barracão | 13 |  | Marmeleiro | 24 |  | Santa Izabel do Oeste |
| 3 |  | Bela Vista da Caroba | 14 |  | Nova Esperança do sudoeste | 25 |  | Santo Antônio do Sudoeste |
| 4 |  | Boa Esperança do Iguaçu | 15 |  | Nova Prata Do Iguaçu | 26 |  | São Jorge D'oeste |
| 5 |  | Bom Jesus do Sul | 16 |  | Pérola D'oeste | 27 |  | Verê |
| 6 |  | Capanema | 17 |  | Pinhal de São Bento | 28 |  | Outro do Paraná  Another municipality of Paraná |
| 7 |  | Cruzeiro do Iguaçu | 18 |  | Planalto | 29 |  | Rio Grande de Sul |
| 8 |  | Dois Vizinhos | 19 |  | Pranchita | 30 |  | Santa Catarina |
| 9 |  | Éneas Marques | 20 |  | Realeza | 31 |  | Outro estado do Brasil  Another state of Brazil |
| 10 |  | Flor da Serra Do Sul | 21 |  | Renascença |  |  |  |
| 11 |  | Francisco Beltrão | 22 |  | Salgado Filho |  |  |  |

2. **Where you live?**

| 1 |  | City | 2 |  | Rural |
| --- | --- | --- | --- | --- | --- |

3. **How many years do you live in this place?**

| 1 |  | 1-5 | 5 |  | 21-25 | 9 |  | 41-45 |
| --- | --- | --- | --- | --- | --- | --- | --- | --- |
| 2 |  | 6-10 | 6 |  | 26-30 | 10 |  | 46-50 |
| 3 |  | 11-15 | 7 |  | 31-35 | 11 |  | Over 51 |
| 4 |  | 16-20 | 8 |  | 36-40 |  |  |  |

**4. Do you have dairy cows?**

| 1 |  | No | 2 |  |  | Yes |
| --- | --- | --- | --- | --- | --- | --- |

4. **Which products your family plants with pesticides?**

| 1  1 |  | Soybean/Corn | 5  5 |  | Tobacco | 9  9 |  | Vegetables |
| --- | --- | --- | --- | --- | --- | --- | --- | --- |
| 2  2 |  | Soybean/Wheat | 6  6 |  | Pasture | 10  10 |  | Others |
| 3  3 |  | Corn/Pasture | 7  7 |  | Beans | 11  11 |  | Fruits and Vegetables |
| 4  4 |  | Corn/Wheat | 8  8 |  | Fruits | 12  12 |  | Does not apply |

5. **Which products do you plant and consume with pesticides?**

| 1  1 |  | Corn | 5  5 |  | Tobacco |
| --- | --- | --- | --- | --- | --- |
| 2  2 |  | Bean | 6  6 |  | Others |
| 3  3 |  | Fruits | 7  7 |  | Fruits and Vegetables |
| 4  4 |  | Vegetables | 8  8 |  | Does not apply |

6. **Do you currently apply any type of pesticides in gardens, flowers or crop? (If the answer is yes, complete BOX 1)**

| 1  1 |  | No | 2  2 |  | Yes |
| --- | --- | --- | --- | --- | --- |

7. **Does your family use pesticides today? (If the answer is yes, complete BOX 1)**

| 1 |  | No | 2 |  | Yes |
| --- | --- | --- | --- | --- | --- |

8. **Who applies these products?**

| 1  1 |  | Father | 3  3 |  | Brothers | 5  5 |  | Husband | 7  7 |  | Don’t know | 9  9 |  | Others |
| --- | --- | --- | --- | --- | --- | --- | --- | --- | --- | --- | --- | --- | --- | --- |
| 2  2 |  | Mother | 4  4 |  | Grandfather | 6  6 |  | Sons | 8  8 |  | Does not apply |  |  |  |

**BOX 1 – EXPOSITION TO PESTICIDES IN THE PRESENT**

| PESTICIDE NAME | CULTURE | PERIOD | | AMOUNT USED | WHO APPLIES |
| --- | --- | --- | --- | --- | --- |
|  | | Month | Number of Days |  | |

9. **Have you ever lived in another municipality?**

| 1  1 |  |  | No | 2  2 |  | Yes |
| --- | --- | --- | --- | --- | --- | --- |

10. **If the answer is yes, which place?**

| 1 |  | Ampére | 12 |  | Manfrinópolis | 23 |  | Salto do Lontra |
| --- | --- | --- | --- | --- | --- | --- | --- | --- |
| 2 |  | Barracão | 13 |  | Marmeleiro | 24 |  | Santa Izabel do Oeste |
| 3 |  | Bela Vista da Caroba | 14 |  | Nova Esperança do sudoeste | 25 |  | Santo Antônio do Sudoeste |
| 4 |  | Boa Esperança do Iguaçu | 15 |  | Nova Prata Do Iguaçu | 26 |  | São Jorge D'oeste |
| 5 |  | Bom Jesus do Sul | 16 |  | Pérola D'oeste | 27 |  | Verê |
| 6 |  | Capanema | 17 |  | Pinhal de São Bento | 28 |  | Another municipality of Paraná state |
| 7 |  | Cruzeiro do Iguaçu | 18 |  | Planalto | 29 |  | Rio Grande de Sul |
| 8 |  | Dois Vizinhos | 19 |  | Pranchita | 30 |  | Santa Catarina |
| 9 |  | Éneas Marques | 20 |  | Realeza | 31 |  | Another state of Brazil |
| 10 |  | Flor da Serra Do Sul | 21 |  | Renascença | 32 |  | Did not live |
| 11 |  | Francisco Beltrão | 22 |  | Salgado Filho |  |  |  |

11. **Have you ever lived in the countryside?**

| 1  1 |  | No | 2  2 |  | Yes |
| --- | --- | --- | --- | --- | --- |

12. **If the answer is yes, how many years?**

| 1 |  | 1-5 | 5 |  | 21-25 | 9 |  | 41-45 |
| --- | --- | --- | --- | --- | --- | --- | --- | --- |
| 2 |  | 6-10 | 6 |  | 26-30 | 10 |  | 46-50 |
| 3 |  | 11-15 | 7 |  | 31-35 | 11 |  | + de 51 |
| 4 |  | 16-20 | 8 |  | 36-40 | 12 |  | Does not apply |

13. **In the places where you lived, your family used pesticides? (If the answer is yes, please complete BOX 2)**

| 1  1 |  | No | 2  2 |  | Yes |
| --- | --- | --- | --- | --- | --- |

14. **Have you ever applied pesticides? (**If the answer is yes, complete Table 2)

| 1  1 |  | No | 2  2 |  | Yes |
| --- | --- | --- | --- | --- | --- |

**BOX** 2 – EXPOSITION TO PESTICIDES IN THE PAST

| PESTICIDE NAME | CULTURE | PERIOD | | AMOUNT USED | WHO APPLIED |
| --- | --- | --- | --- | --- | --- |
|  | | Month | Number of Days |  | |

15. **How many years have you applied pesticides?**

| 1 |  | 1-5 | 5 |  | 11-25 | 9 |  | 41-45 |
| --- | --- | --- | --- | --- | --- | --- | --- | --- |
| 2 |  | 6-10 | 6 |  | 26-30 | 10 |  | 46-50 |
| 3 |  | 11-15 | 7 |  | 31-35 | 11 |  | Over 51 |
| 4 |  | 16-20 | 8 |  | 36-40 | 12 |  | Does not apply |

16. **Who in your family also applied pesticides?**

| 1  1 |  | Father | 3  3 |  | Brother/Sister | 5  5 |  | Husband | 7  7 |  | Do not know |
| --- | --- | --- | --- | --- | --- | --- | --- | --- | --- | --- | --- |
| 2  2 |  | Mother | 4  4 |  | Grandfather/Grandmother | 6  6 |  | Son/Daughter | 8  8 |  | Does not apply |

17. **For how many years?**

| 1 |  | 1-5 | 5 |  | 21-25 | 9 |  | 41-45 |
| --- | --- | --- | --- | --- | --- | --- | --- | --- |
| 2 |  | 6-10 | 6 |  | 26-30 | 10 |  | 46-50 |
| 3 |  | 11-15 | 7 |  | 31-35 | 11 |  | Over 51 |
| 4 |  | 16-20 | 8 |  | 36-40 | 12 |  | Does not apply |

18. **Do you wash any family's clothes after the application of pesticides?**

| 1 |  | No | 2 |  | Yes | 3 |  | Does not apply |
| --- | --- | --- | --- | --- | --- | --- | --- | --- |

19. **Did you wash or washed any family's clothes after the pesticide’s application?**

| 1 |  | No | 2 |  | Yes | 3 |  | Does not apply |
| --- | --- | --- | --- | --- | --- | --- | --- | --- |

20. **Do you wear or did you wear gloves to wash these clothes?**

| 1 |  | No | 2 |  | Yes | 3 |  | Does not apply |
| --- | --- | --- | --- | --- | --- | --- | --- | --- |

21. **Do you wash or did you wash these contaminated clothes along with other family clothes?**

| 1 |  | No | 2 |  | Yes | 3 |  | Does not apply |
| --- | --- | --- | --- | --- | --- | --- | --- | --- |

22. **Do any neighbor of yours use pesticides?**

| 1 |  | No | 2 |  | Yes | 3 |  | Does not apply |
| --- | --- | --- | --- | --- | --- | --- | --- | --- |

23. **Did any neighbor of yours use pesticides?**

| 1 |  | No | 2 |  | Yes | 3 |  | Does not apply |
| --- | --- | --- | --- | --- | --- | --- | --- | --- |

24. **Do you use or use protective equipment when using the pesticide?**

| 1 |  | No | 2 |  | Yes | 3 |  | Does not apply |
| --- | --- | --- | --- | --- | --- | --- | --- | --- |

25. **Do Other people in your family use or use protective equipment when using the pesticide?**

| 1 |  | Não  No | 2 |  | Sim  Yes | 3 |  | Não se aplica  Does not apply it |
| --- | --- | --- | --- | --- | --- | --- | --- | --- |

26. **If the answer is yes, which one?**

| 1  1 |  | EVERYONE | 6  6 |  | GLASSES |
| --- | --- | --- | --- | --- | --- |
| 2  2 |  | GLOVES | 7  7 |  | MASK |
| 3  3 |  | BOOTS | 8  8 |  | OTHERS |
| 4  4 |  | GLOVES/BOOTS/MASK | 9  9 |  | Does not apply |
| 5  5 |  | Overalls |  |  |  |

27. **Did you notice any changes (smell, dizziness, headache, etc.) when you had direct or indirect contact with pesticides?**

| 1 |  | No | 2 |  | Yes | 3 |  | Does not apply |
| --- | --- | --- | --- | --- | --- | --- | --- | --- |

**If the answer is yes, which one? _______________________________________________________________________________**

28. **Have your relatives had these symptoms?**

**If the answer is yes, which one? ___________________________________________________________________________**

29. **Did you feed on what was produces on your property with pesticide?**

| 1 |  | No | 2 |  | Yes | 3 |  | Does not apply it |
| --- | --- | --- | --- | --- | --- | --- | --- | --- |

**If the answer is yes, which one?**

| 1  1 |  | Corn | 5  5 |  | Tobacco |
| --- | --- | --- | --- | --- | --- |
| 2  2 |  | Bean | 6  6 |  | Others |
| 3  3 |  | Fruits | 7  7 |  | Fruits and Vegetables |
| 4  4 |  | Vegetables | 8  8 |  | Does not apply |

30. **Where does drink water come from?**

| 1  1 |  | Private well | 4  4 |  | Network |
| --- | --- | --- | --- | --- | --- |
| 2  2 |  | Community well | 5  5 |  | River |
| 3  3 |  | Source | 6  6 |  | Mineral water |

31. **How is the distance from this water to tillage areas?**

| 1 |  | Till 5 meters | 4 |  | 21 a 30 m |
| --- | --- | --- | --- | --- | --- |
| 2 |  | 6 a 10 m | 5 |  | More than 31 m |
| 3 |  | 11 a 20 m | 6 |  | Does not apply |

**HEALTH AND INTOXICATION**

32. **Have you ever had any pesticide intoxication?**

| 1  1 |  | No | 2  2 |  | Yes |
| --- | --- | --- | --- | --- | --- |

33. **How many times?**

| 1 |  | Once | 4 |  | 4 Times | 7 |  | Never got intoxicated |
| --- | --- | --- | --- | --- | --- | --- | --- | --- |
| 2 |  | Twice | 5 |  | 5 Times |  |  |  |
| 3 |  | 3 Times | 6 |  | More than 5 times |  |  |  |

34. **Which symptoms did you have?**

| 1  1 |  | Gastrointestinals | 6  6 |  | Do not remember |
| --- | --- | --- | --- | --- | --- |
| 2  2 |  | Sensory/neurological alteration | 7  7 |  | Other, please describe which ones: |
| 3  3 |  | Skin alteration | 8  8 |  | Gastrointestinal, skin and respiratory disorders |
| 4  4 |  | Cardiovascular alteration | 9  9 |  | Does not apply |
| 5  5 |  | Respiratory Alteration |  |  |  |

35. **Location where it was attended:**

| 1  1 |  | Hospital | 4  4 |  | Private physician’s office |
| --- | --- | --- | --- | --- | --- |
| 2  2 |  | Health unit | 5  5 |  | Did not seek health care |
| 3  3 |  | Emergency and Urgency Center | 6  6 |  | Does not apply |

36. **Has anyone in your family ever had pesticides intoxication?**

| 1  1 |  | No | 2  2 |  | Yes | 3  3 |  | Do not you know |
| --- | --- | --- | --- | --- | --- | --- | --- | --- |

37. **Who?**

| 1  1 |  | Father | 3  3 |  | Brother/Sister | 5  5 |  | Husband | 7  7 |  | Others | 9  9 |  | Does not apply |
| --- | --- | --- | --- | --- | --- | --- | --- | --- | --- | --- | --- | --- | --- | --- |
| 2  2 |  | Mother | 4  4 |  | Grandfather/grandmother | 6  6 |  | Son/Daughter | 8  8 |  | Do not know |  |  |  |

38. **How many times?**

| 1  1 |  | Once | 4  4 |  | 4 times | 7  7 |  | Does not apply |
| --- | --- | --- | --- | --- | --- | --- | --- | --- |
| 2  2 |  | Twice | 5  5 |  | 5 times | 8  8 |  |  |
| 3  3 |  | 3 times | 6  6 |  | More than 5 times | 9  9 |  |  |

39. **Which symptoms did you have?**

| 1  1 |  | Gastrointestinals | 6  6 |  | Do not remember |
| --- | --- | --- | --- | --- | --- |
| 2  2 |  | Sensory/neurological alteration | 7  7 |  | Other, please describe which ones: |
| 3  3 |  | Skin alteration | 8  8 |  | Gastrointestinal, skin and respiratory disorders |
| 4  4 |  | Cardiovascular alteration | 9  9 |  | Does not apply |
| 5  5 |  | Respiratory Alteration |  |  |  |

40. **Location where it was attended**

| 1  1 |  | Hospital | 4  4 |  | Private physician’s office |
| --- | --- | --- | --- | --- | --- |
| 2  2 |  | Health unit | 5  5 |  | Did not seek health care |
| 3  3 |  | Emergency and Urgency Center | 6  6 |  | Does not apply |

41. **Do you have any disease?**

| 1  1 |  | No | 2  2 |  | Yes |
| --- | --- | --- | --- | --- | --- |

42. **If the answer is yes, which one(s]?**

__________________________________________________________________________________________________________________________________________________________________________________________________________________________________________________________________________________________________________________________________________________________________

43. **Does anyone in your family have any disease? If the answer is yes, complete BOX 3**

| 1  1 |  | No | 2  2 |  | Yes |
| --- | --- | --- | --- | --- | --- |

**BOX 3**

| Family member ID | Disease | How long? |
| --- | --- | --- |
|  |  | \| 0 to 1 year \| \| 2 to 3 years; \| 4 to 5 years \| \| More than 5 years |
|  |  | \| 0 to 1 year \| \| 2 to 3 years; \| 4 to 5 years \| \| More than 5 years |

44. **Have you ever had breast cancer or any other type of cancer?**

| 1  1 |  | No | 2  2 |  | Yes |
| --- | --- | --- | --- | --- | --- |

**If the answer is yes, which one?** ________________________________________________________________________

45. **Does anyone in your family ever had cancer? If the answer is yes, complete BOX 4**

| 1  1 |  | No | 2  2 |  | Yes |
| --- | --- | --- | --- | --- | --- |

**BOX 4**

| Family member | Type of cancer | Overcome |
| --- | --- | --- |
|  |  | ( ) cure / ( ) sequel / ( ) death |
|  |  | ( ) cure / ( ) sequel / ( ) death |
|  |  | ( ) cure / ( ) sequel / ( ) death |

46. **Have you ever had loss of movement, tingling, blurred vision, loss of muscle strength or change in sensitivity?**

| 1  1 |  | No | 2  2 |  | Yes |
| --- | --- | --- | --- | --- | --- |

47. **If the answer is yes, which problem? __**_________________________________________________________
48. **Were you born with any malformation?**

| 1  1 |  | No | 2  2 |  | Yes |
| --- | --- | --- | --- | --- | --- |

49. **If the answer is yes, which one?** _______________________________________________________________________
50. **AWas anyone in your family born with any malformation? If the answer is yes, complete BOX 5**

| 1  1 |  | No | 2  2 |  | Yes | 3  3 |  | Do not know |
| --- | --- | --- | --- | --- | --- | --- | --- | --- |

**BOX 5**

| Family member | Type of malformation |
| --- | --- |

51. **Have you ever had a miscarriage?**

| 1  1 |  | Não  No | 2  2 |  | Sim  Yes |
| --- | --- | --- | --- | --- | --- |

52. **Has anyone in your family ever had a miscarriage?**

| 1  1 |  | No | 2  2 |  | Yes | 3  3 |  | Do not know |
| --- | --- | --- | --- | --- | --- | --- | --- | --- |

53. **Have you ever had any difficulty to getting pregnant?**

| 1  1 |  | No | 2  2 |  | Yes |
| --- | --- | --- | --- | --- | --- |

54. **Did anyone in your Family have any trouble getting pregnant?**

| 1  1 |  | No | 2  2 |  | Mother | 3  3 |  | Sister | 4  4 |  | Daughter | 5  5 |  | Grandmother | 6  6 |  | Others | 7  7 |  | Do not know |
| --- | --- | --- | --- | --- | --- | --- | --- | --- | --- | --- | --- | --- | --- | --- | --- | --- | --- | --- | --- | --- |

55. **Have you been exposed to pesticide during your children’s pregnancy?**

| 1  1 |  | No | 2  2 |  | Yes |
| --- | --- | --- | --- | --- | --- |

56. **Was the husband exposed to pesticide before the children were conceptioned?**

| 1  1 |  | No | 2  2 |  | Yes |
| --- | --- | --- | --- | --- | --- |

57. **Have you ever done blood tests in your life to assess exposure to pesticide (cholinesterase)?**

| 1  1 |  | No | 2  2 |  | Yes |
| --- | --- | --- | --- | --- | --- |

58. **Has anyone in your family ever done blood tests in their lives to assess exposure to pesticide (cholinesterase)?**

| 1  1 |  | No | 2  2 |  | Yes |
| --- | --- | --- | --- | --- | --- |
